# Unique Behavior Profiles that Specify Mental Distress in Autism

**DOI:** 10.1101/2025.02.01.25321517

**Authors:** Eric V. Strobl

**Affiliations:** University of Pittsburgh

## Abstract

**Objective:** Patients with autism spectrum disorder (ASD) often have difficulty describing their emotions, leaving clinicians to infer mental distress from aberrant behaviors. Unfortunately, aberrant behaviors are non-specific and can differ from those of typically developing peers. However, aberrant behavior profiles (ABPs), or weighted combinations of *multiple* aberrant behaviors, may be specific and recognizable. We thus sought to identify ABPs that correlate with distinct types of mental distress, and also correlate differently in ASD than in typical development (TD).

**Method:** We integrated three studies that measured diagnostic symptom severity using the Child and Adolescent Symptom Inventory, and aberrant behaviors using the Aberrant Behavior Checklist. We then performed a component analysis to identify maximally non-overlapping severity scores that correlate with ABPs differently in ASD than in TD.

**Results:** Behaviors associated with impulsivity and distractibility correlated more positively with the severity of specific phobia and social anxiety in ASD than in TD. Hyperactivity and tantrums correlated more positively with attention-deficit hyperactivity disorder and oppositional defiant disorder in ASD than in TD. Emotional reactivity correlated more positively with obsessions and somatization, but social withdrawal and self-injurious behaviors correlated more negatively; the opposite pattern held in a second phenotype of social anxiety. Finally, preoccupation correlated positively with schizophrenia, but depression and inactivity correlated negatively; the opposite pattern held with the severity of post-traumatic stress disorder and a second phenotype of specific phobia.

**Conclusion:** Patients with ASD (1) externalize more in attention-deficit hyperactivity disorder and oppositional defiant disorder, (2) exhibit unique signs of internal distress in post-traumatic stress and psychosis, and (3) display highly distinct ABPs in specific forms of anxiety.

## Introduction

Patients with autism spectrum disorder (ASD) often have difficulty with emotional self-awareness^1^ and exhibit a higher frequency of externalizing behaviors, especially among children and adolescents, as compared to their typically developing peers.^2^ As a result, caregivers of patients with ASD frequently only report non-specific aberrant behaviors that appear to emerge spontaneously. For example, a patient may suddenly throw tantrums, cry incessantly and bang their head on objects for reasons unknown to the caregiver. This places caregivers and clinicians in the difficult position of needing to deduce the underlying mental distress based solely on a set of observable behaviors.

Inferring internal distress from observable behaviors carries multiple challenges. First, many aberrant behaviors, such as tantrums or body rocking, are not specific to any single type of mental distress; an aberrant behavior can only suggest particular types of mental distress with a high degree of uncertainty.^3^ Second, patients with ASD exhibit behaviors that can differ from typically developing peers even when under the same type of mental distress.^4^ For example, patients with ASD often engage in self-injury in social situations due to sensory overload and anxiety,^5^ whereas typically developing peers tend to injure themselves when experiencing post-traumatic stress or depression.^6^ Caregivers and clinicians can thus have trouble accurately inferring the mental distress of a patient with ASD. We need evidence-based guidelines that can help translate aberrant behaviors into psychiatric severity scores to accurately deduce mental distress in ASD.

We cannot ask all patients with ASD how they feel and obtain detailed answers. Many patients also do not meet strict diagnostic cutoffs for particular comorbid disorders. We thus indirectly inferred mental distress by adopting the strategy in Figure 1, where we correlated *highly specific behaviors* – known to predict the ordinal severity levels of distinct types of mental distress in children and adolescents – to various *non-specific aberrant behaviors*. Note that the most challenging aberrant behaviors to interpret are those that correlate with mental distress differently in patients with ASD than in typically developing peers. We therefore further identified distinct sets of non-specific aberrant behaviors, or Aberrant Behavior Profiles (ABPs), that correlate with specific types of mental distress differently in ASD. We hypothesized that patients with ASD exhibit different but recognizable ABPs indicative of specific types of mental distress.

**Figure 1.**
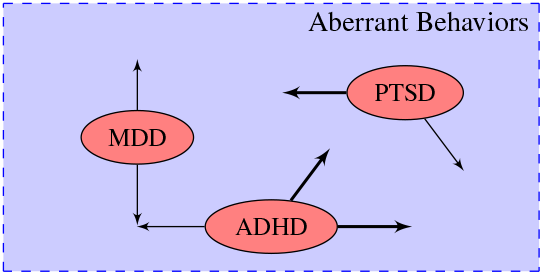
Strategy overview. To infer mental distress in even non-verbal patients, we correlate highly specific behaviors indicative of the severity level of certain diagnostic categories (red ellipses) to various aberrant behaviors (blue rectangle) in children and adolescents. Patients thus do not need to meet strict diagnostic cutoff scores but only show *some* signs of comorbid mental illnesses. We seek to discover the aberrant behaviors denoted by the thick arrows that (1) correlate differently in patients with ASD than in TD, and (2) maximally specify particular diagnostic categories. MDD denotes major depressive disorder, ADHD attention-deficit hyperactivity disorder, PTSD post-traumatic stress disorder.

## Method

### Rating Scales

The following rating scales quantify the diagnostic severity scores and aberrant behaviors needed to implement the strategy summarized in Figure 1:

1. Child and Adolescent Symptom Inventory, version 5 (CASI-5):^7^ this scale evaluates behaviors that specify particular emotional and behavioral disorders in children and adolescents according to the Diagnostic and Statistical Manual of mental disorders version 5 (DSM-5). A caregiver rates each behavior based on its degree of frequency: 0 = never; 1 = sometimes; 2 = often; 3 = very often. We created severity scores by summing the frequency scores of each behavior assigned to one of 12 common diagnostic categories associated with internal distress: ADHD inattentive type, ADHD hyperactive type, oppositional defiant disorder (ODD), generalized anxiety disorder (GAD), specific phobia, obsessions, PTSD, somatization, social anxiety, separation anxiety, schizophrenia, and MDD.
2. Aberrant Behavior Checklist, community version (ABC):^8, 9^ this scale contains 58 problematic or aberrant behaviors rated as: 0 = not at all a problem; 1 = the behavior is a problem but slight in degree; 2 = the problem is moderately serious; 3 = the problem is severe in degree. Investigators originally designed ABC for patients with intellectual disability living in institutional or residential facilities. A later community version extended ABC to home and school settings. Exploratory factor analyses on the items have consistently revealed a five-factor structure across multiple populations, including children and adolescents with ASD.^10^

CASI-5 thus focuses on specific behaviors associated with common diagnostic categories of the DSM-5, whereas ABC includes a variety of mostly non-specific aberrant behaviors.

### Datasets

We searched the entire National Institute of Mental Health (NIMH) Data Archive using the keyword “autism.” The search yielded 644 datasets on January 2nd, 2025. We then manually included any dataset containing at least one patient with ASD and complete data for all of the following variables in at least one time point: age, biological sex, Autism Diagnostic Observation Schedule Second Edition (ADOS-2) module numbers and diagnoses,^11^ all individual ABC items, all individual CASI-5 items. We excluded datasets that only recruited patients with ASD and specific psychiatric comorbidities, such as anxiety, to avoid selection bias on the CASI-5 severity scores. Three datasets met the inclusion and exclusion criteria:

**Table 1:** Summary statistics of the three datasets meeting study inclusion and exclusion criteria. Dataset ID refers to the unique NIMH Data Archive ID. SD denotes standard deviation. We present detailed summary statistics across all three datasets in Results.

| Dataset ID | Ref. | ASD<br>no. | TD<br>no. | Age<br>mean (SD) | Sex<br>% male | Race<br>% white |
| --- | --- | --- | --- | --- | --- | --- |
| 3670 | [12] | 151 | 81 | 12.7 (1.85) | 75.4 | 73.2 |
| 2025 | [13] | 83 | 22 | 13.1 (4.22) | 72.3 | 65.1 |
| 2181 | [14] | 9 | 0 | 6.20 (1.29) | 66.7 | 77.8 |

We downloaded the above individual-level datasets from the NIMH Data Archive using a limited access data use certificate awarded to author ### (anonymized).

### Dependent, Independent, Nuisance and Moderator Variables

We seek to identify CASI-5 severity scores that correlate with the ABC aberrant behaviors differently in patients with ASD than in typically developing peers. We therefore set the CASI-5 severity scores as the independent variables, and the ABC items as the dependent variables. We sought ABPs invariant to age and biological sex, so we set these two variables as nuisance variables and partialed them out using ordinary least squares before performing downstream analyses. Finally, patients with minimally verbal ASD (MV-ASD) may exhibit different ABPs than patients with fluently verbal ASD (FV-ASD). We thus set the verbal fluency status of a patient with ASD as a moderator variable. In particular, we categorized patients as FV-ASD if they could complete ADOS-2 module 3 or 4 with a resultant ASD diagnosis.^11^ We categorized patients as MV-ASD if they could only complete ADOS-2 module 1 or 2 with a resultant ASD diagnosis.

### Learning Aberrant Behavior Profiles

We created the Differentially Supervised Varimax (DSV) algorithm to learn conceptually distinct ABPs that correlate with non-overlapping diagnostic severity scores differently in ASD than in TD. The DSV algorithm is a derivative of the Supervised Varimax algorithm originally proposed in [15] but modified to directly accomplish the task proposed in this paper. DSV first combines the individual items in the ABC scale ***Y*** into *m* orthogonal, and thus conceptually distinct, principal components ***C***. The CASI-5 scores ***X*** are correlated to many components ***C*** differently in patients with ASD than in TD. The resultant model is thus dense with many non-zero differential correlations from ***X*** to ***C***, and many non-zero effect sizes from ***C*** to ***Y*** (Figure 2 (a)).

**Figure 2.**
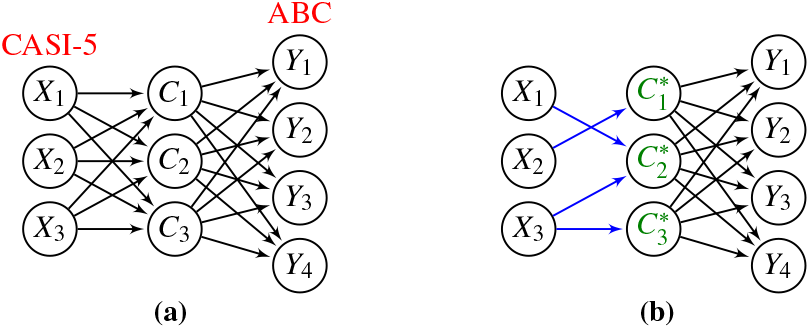
Overview of the DSV algorithm. (a) DSV considers a model where CASI-5 severity scores ***X*** are correlated to the orthogonal components ***C*** differently in ASD than in TD. The components in turn influence ABC aberrant behaviors ***Y*** . Unfortunately, the model is complex due to many differential correlations between ***X*** and ***C***. (b) DSV simplifies the model by rotating the components into a set of optimal but still orthogonal components ***C***^∗^ such that the differential correlations from ***X*** and ***C***^∗^ are now sparse. As a result, only a few non-overlapping severity scores are differentially correlated with each optimal component encapsulating an orthogonal, or conceptually distinct, combination of aberrant behaviors.

DSV rotates the components into a set of optimal but still orthogonal components ***C***^∗^ using the varimax rotation^16^ such that each optimal component is correlated to only a few non-overlapping CASI-5 scores differently in ASD than in TD (Figure 2 (b)). For example, severity score *X*_1_ differentially correlates with all three optimal components in Figure 2 (a), but *X*_1_ only differentially correlates with the second optimal component in Figure 2 (b). As a result, only a few non-overlapping severity scores are differentially correlated with each conceptually distinct ABP in Figure 2 (b), as desired. We provide a detailed technical description of DSV in the Supplementary Material. We learned *m* = 10 optimal components due to a steep cliff observed in the scree plot after the tenth eigenvalue (Supplementary Material Figure S1).

### Hypothesis Testing

We performed two-tailed permutation testing by component and by severity-component pairs all under the null hypothesis of exchangeability of the ASD and TD diagnoses. The alternative hypothesis corresponds to a differential correlation in a specific optimal component or specific severity-optimal component pair. We corrected for multiple comparisons by controlling the false discovery rate (FDR). We always performed 100,000 permutations. We provide additional descriptions of each permutation test in the Supplementary Material.

## Results

### Patient Characteristics

We summarize patient characteristics after combining all three datasets in Table 2 with associated *p*-values. Subjects with ASD and TD had approximately the same mean age and sex ratio. Dataset 2025 had no racial information for subjects with TD. Subjects of Asian descent were more likely to participate in the study if they had a diagnosis of ASD, especially MV-ASD. No significant difference between racial groups existed between ASD and TD (*p*=0.321), and between FV-ASD and MV-ASD (*p*=0.859), when we removed Asian subjects. Patients with ASD had more frequent and severe aberrant behaviors according to all subscales of the ABC. Patients similarly had more frequent and severe symptoms according to the CASI-5 severity scores. The differences between patients with FV-ASD and MV-ASD were smaller, but most of the *p*-values still fell below the uncorrected 5% threshold.

**Table 2:** Summary of patient characteristics after combining the three datasets. One of the datasets had incomplete racial data, so we only present the counts for the complete cases. Continuous values are summarized by their mean and, in parentheses, standard deviation. We report *p*-values of two-sample two-sided *t* -tests for continuous variables, and *p*-values of Fisher’s exact test for counts (biological sex, race). Subjects with ASD and TD showed marked differences in all ABC subscales and CASI-5 severity scores. Subjects with FV-ASD and MV-ASD also had many differences, but the differences were smaller.

|  | ASD<br>(*n*=243) | TD<br>(*n*=103) | ASD vs. TD<br>*p*-value | FV-ASD<br>(*n*=156) | MV-ASD<br>(*n*=87) | FV vs. MV<br>*p*-value |
| --- | --- | --- | --- | --- | --- | --- |
| Demographics |  |  |  |  |  |  |
| Age | 12.4 (3.18) | 13.0 (2.37) | 0.056 | 13.1 (2.62) | 11.2 (3.72) | <0.001 |
| Biological sex |  |  |  |  |  |  |
| Male | 184 | 73 | 0.350 | 121 | 63 | 0.436 |
| Female | 59 | 30 |  | 35 | 24 |  |
| Race (complete) |  |  |  |  |  |  |
| White | 164 | 67 | 0.002 | 113 | 51 | 0.001 |
| Black | 11 | 4 |  | 8 | 3 |  |
| Asian | 25 | 0 |  | 7 | 18 |  |
| Other | 43 | 10 |  | 28 | 15 |  |
| ABC subscales |  |  |  |  |  |  |
| Irritability | 15.0 (9.29) | 3.60 (4.10) | <0.001 | 15.5 (9.45) | 14.1 (8.98) | 0.256 |
| Lethargy | 31.3 (14.8) | 12.1 (10.4) | <0.001 | 31.5 (14.9) | 30.9 (14.8) | 0.762 |
| Stereotypy | 3.98 (3.07) | 0.30 (1.03) | <0.001 | 3.61 (2.99) | 4.66 (3.09) | 0.011 |
| Hyperactivity | 22.1 (13.9) | 5.27 (8.23) | <0.001 | 20.7 (13.6) | 24.5 (14.1) | 0.046 |
| Inappropriate speech | 3.07 (2.34) | 0.73 (1.38) | <0.001 | 2.63 (2.10) | 3.85 (2.55) | <0.001 |
| CASI-5 severity scores |  |  |  |  |  |  |
| ADHD inattention | 14.7 (6.26) | 6.12 (5.05) | <0.001 | 14.9 (6.18) | 14.5 (6.44) | 0.692 |
| ADHD hyperactivity | 10.9 (6.77) | 2.93 (4.75) | <0.001 | 10.2 (6.83) | 12.1 (6.52) | 0.032 |
| ODD | 6.81 (5.57) | 4.49 (3.92) | <0.001 | 7.99 (5.73) | 4.69 (4.58) | <0.001 |
| GAD | 5.30 (3.90) | 2.07 (2.13) | <0.001 | 6.29 (3.90) | 3.53 (3.24) | <0.001 |
| Specific phobia | 1.06 (1.20) | 0.30 (0.64) | <0.001 | 1.22 (1.29) | 0.77 (0.96) | 0.002 |
| Obsessions | 0.63 (0.85) | 0.20 (0.47) | <0.001 | 0.84 (0.90) | 0.24 (0.57) | <0.001 |
| PTSD | 0.83 (1.30) | 0.11 (0.42) | <0.001 | 0.99 (1.41) | 0.54 (1.04) | 0.005 |
| Somatization | 0.74 (1.10) | 0.36 (0.79) | <0.001 | 1.04 (1.17) | 0.21 (0.68) | <0.001 |
| Social anxiety | 3.48 (3.07) | 0.71 (1.88) | <0.001 | 3.69 (3.08) | 3.10 (3.04) | 0.156 |
| Separation anxiety | 1.99 (3.37) | 0.48 (1.20) | <0.001 | 2.17 (3.76) | 1.67 (2.53) | 0.212 |
| Schizophrenia | 2.42 (2.65) | 0.20 (0.62) | <0.001 | 2.36 (2.70) | 2.54 (2.55) | 0.604 |
| MDD | 3.35 (4.21) | 1.96 (2.73) | <0.001 | 4.51 (4.55) | 1.28 (2.40) | <0.001 |

### ASD versus TD

The DSV algorithm discovered 4 significant optimal components at an FDR threshold of 5%. These optimal components exhibited significant differential correlations with 8 diagnostic categories (Figure 3 red bars). We plot a heat map of the effect sizes from each of the significant optimal components to the 58 aberrant behaviors in the ABC in Figure 4. The 4 optimal components harbored 6 recognizable themes, or ABPs, in the heat map linked to the 8 diagnostic severity scores:

**Figure 3.**
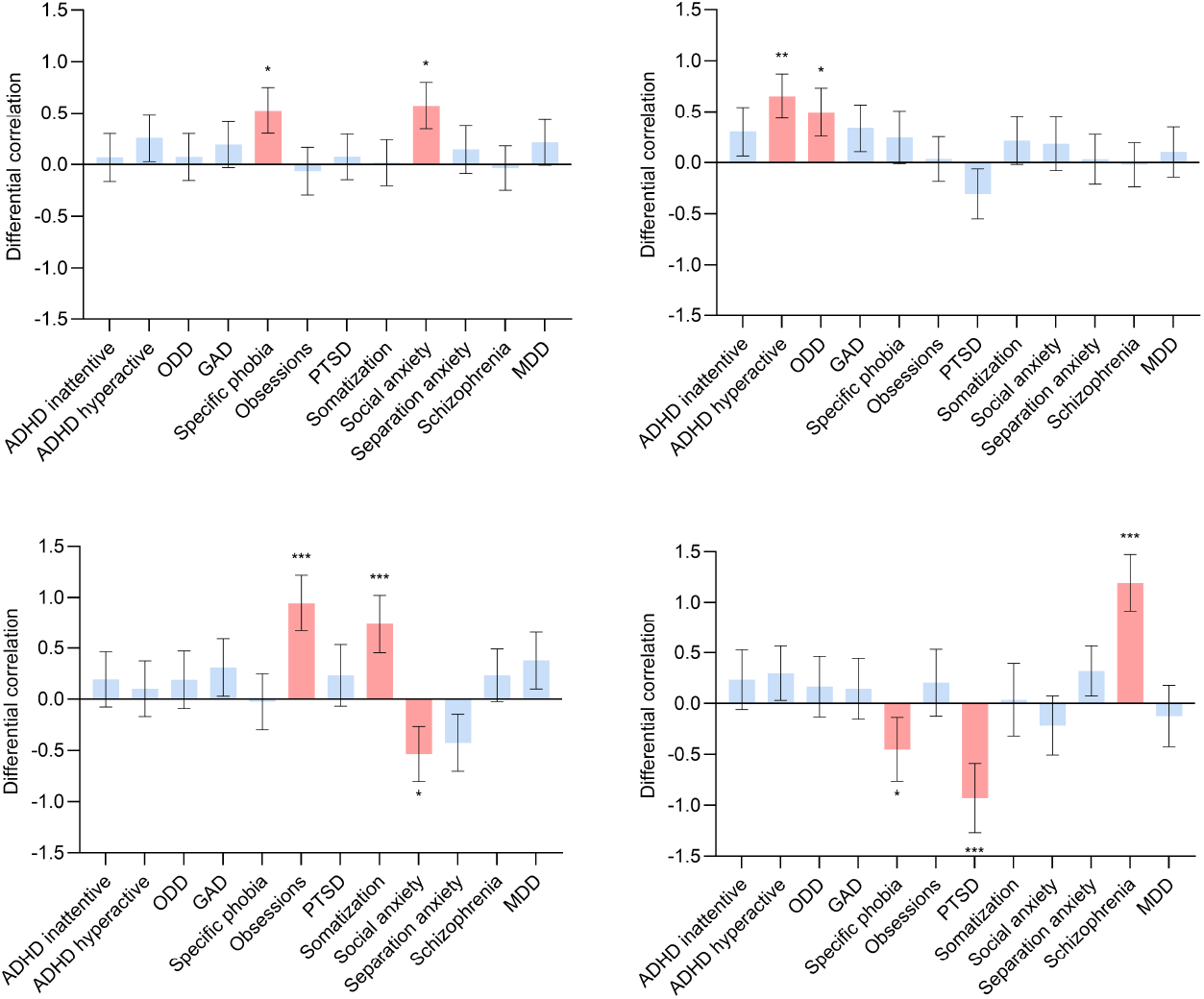
The difference between the correlations of the severity scores to the 4 significant optimal components in ASD vs. TD (blue edges between ***X*** and ***C***^∗^ in Figure 2 (b)). Bars in red highlight severity scores that achieved an FDR below the 5% threshold according to two-sided permutation tests by severity-component. Error bars always denote 95% confidence intervals of the mean. * *p*_*FDR*_ < 0.05, ** *p*_*FDR*_ < 0.01, *** *p*_*FDR*_ < 0.001.

**Figure 4.**
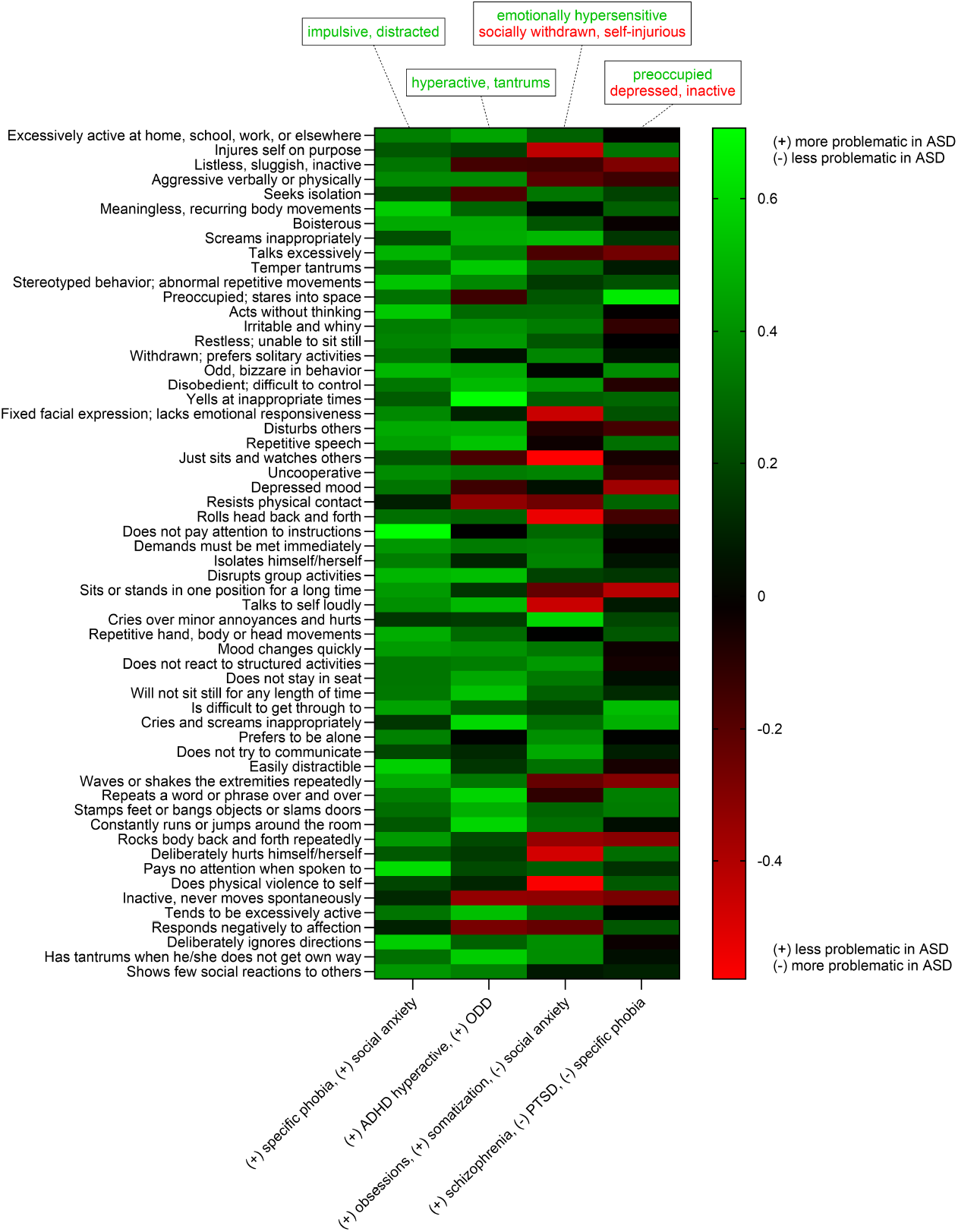
Heatmap of the effect sizes between all aberrant behaviors of the ABC on the y-axis and the 4 significant optimal components on the x-axis (black edges between ***C***^∗^ and ***Y*** in Figure 2 (b)). We have labeled each significant optimal component by the diagnoses with significant differential correlations per Figure 3. The (+) and (-) signs for each diagnosis on the x-axis indicate whether the differential correlations in Figure 3 are positive or negative. As a result, green cells are more problematic in ASD than in TD for diagnoses with a (+) label, and red cells are more problematic in ASD for diagnoses with a (-) label. The boxed phrases on top of the columns summarize the behaviors in green and/or red in each column.

1. Behaviors associated with **impulsivity and distractibility** correlated more positively to the severity of specific phobia and social anxiety in ASD than in TD (Figure 3 (a), Figure 4 column 1; absolute sum = 2.25, *p*_*FDR*_ = 0.035).
2. Behaviors associated with **hyperactivity and tantrums** correlated more positively to the severity of attention-deficit hyperactivity disorder and oppositional defiant disorder in ASD than in TD (Figure 3 (b), Figure 4 column 2; absolute sum = 2.94, *p*_*FDR*_ = 0.005).
3,4. Behaviors associated **emotional hypersensitivity**, such as screaming or crying over minor annoyances, correlated more positively with the severity of obsessions and somatization in ASD than in TD, but **social withdrawal and self-injurious behaviors** correlated more negatively; the opposite pattern held true in social anxiety (Figure 3 (c), Figure 4 column 3; absolute sum = 4.31, *p*_*FDR*_ < 0.001).
5,6. Behaviors associated with **preoccupation** correlated more positively with the severity of schizophrenia in ASD than in TD, but **depression and inactivity** correlated more negatively; the opposite pattern held true in PTSD and specific phobia (Figure 3 (d), Figure 4 column 4; absolute sum = 4.32, *p*_*FDR*_ < 0.001).

We conclude that interpretable ABPs correlated more positively or negatively to specific diagnostic severity scores in ASD versus TD. Moreover, most ABPs associated with one severity score, with the exceptions of social anxiety and specific phobia.

### MV-ASD vs. FV-ASD

We compared MV-ASD and FV-ASD using the DSV algorithm. None of the optimal components differentiated patients with MV-ASD and those with FV-ASD at even trend level after correcting for multiple comparisons (max absolute sum = 2.34, min *p*_*FDR*_ = 0.169). We conclude that DSV did not detect any ABPs that differentiated MV-ASD from FV-ASD with the three datasets.

## Discussion

We developed a statistical approach to identify diagnostic severity scores that correlate with ABPs differently in patients with ASD than in typically developing peers. We found 6 ABPs that correlated differentially with the severity scores of 8 diagnostic categories. Each ABP corresponded to an interpretable combination of multiple aberrant behaviors. Compared to typically developing peers, we found that patients with ASD exhibit (1) increased impulsivity and distractibility in specific phobia and social anxiety, (2) increased hyperactivity and tantrums in ADHD and ODD, increased emotional hypersensitivity but decreased social withdrawal and self-injurious behaviors in obsessions and somatization, (4) increased preoccupations but decreased depression and inactivity in schizophrenia. On the other hand, patients with ASD were also more prone to (5) depression and inactivity in PTSD and specific phobia, as well as (6) social withdrawal and self-injurious behaviors in social anxiety. We conclude that many ABPs correlate with diagnostic severity scores in ASD and differ substantially from those of typically developing peers.

We obtained both expected and illuminating results. As anticipated, patients with ASD exhibit more externalizing behaviors with increasing hyperactivity and anxiety. ADHD, ODD and anxiety, particularly specific phobia and social anxiety, are highly comorbid with ASD.^17, 18^ However, we also found that patients with ASD show clear signs of internalization with a depressed mood and psychomotor inactivity in post-traumatic stress.^19^ Moreover, psychotic symptoms correlate more positively with preoccupation in patients with ASD. Patients with ASD can thus display certain behaviors even more indicative of internalization than typically developing peers.

We importantly found that patients with ASD display highly distinct patterns of anxiety-related behaviors. Patients exhibit increased emotional hypersensitivity to obsessions and somatization but, notably, not to generalized anxiety. Furthermore, patients with ASD are more likely to engage in self-injury in the context of social anxiety rather than to PTSD or OCD, as more commonly observed in typically developing peers.^20^ Finally, a significant portion of patients with ASD exhibit a depressed mood with specific phobias, even though generalized anxiety is more comorbid with major depression in TD.^21^ Patients with ASD thus display different behaviors in specific, rather than generalized, forms of anxiety, possibly secondary to increased cognitive rigidity and restricted interests.

Note that confounding factors may explain the differential correlations listed above, so we cannot infer causality from them.^22^ For example, we found that patients with ASD who also experience obsessions or somatization scream or cry more often in response to minor annoyances. However, the availability of caregivers can also influence these behaviors, since patients with ASD receive more specialized care on average than their typically developing peers.^23^ Patients may therefore scream or cry more often due to the increased availability of adults rather than due to obsessions or somatization. On the other hand, screaming and crying are non-specific behaviors that do not pinpoint a single comorbid diagnosis. As a result, detecting ABPs that specify diagnostic categories remains an important problem in its own right, regardless of whether the differential correlations are causal.

We did not detect any ABPs differentiating patients with MV-ASD and FV-ASD. Patients with MV-ASD and FV-ASD share many similarities, so detecting differences between MV-ASD and FV-ASD is likely harder than detecting differences between ASD and TD.^24^ Moreover, our sample size of FV-ASD (156 patients) and MV-ASD (87 patients) was smaller than the broader groups of ASD (243 patients) and TD (103 patients). We thus had smaller sample sizes for a harder problem, limiting our ability to draw definitive conclusions about the differences between MV-ASD and FV-ASD.

The DSV algorithm uses a fixed set of diagnostic severity scores, but an alternative set of diagnoses may offer a better categorization of mental distress in ASD.^25^ For example, the CASI-5 severity scores do not account for cognitive rigidity in ASD, even though cognitive rigidity may underlie many of the ABPs related to anxiety. Future work should consider developing a tailored set of severity scores in patients with ASD, rather than relying on a fixed set of categories established for the general population.

In conclusion, we developed a customized component analysis algorithm that identified multiple ABPs correlating with specific diagnostic severity scores differently in ASD than in TD. We found that patients with ASD (1) externalize more in ADHD and ODD, (2) exhibit unique signs of internal distress in post-traumatic stress and psychosis, and (3) display highly distinct ABPs in specific forms of anxiety. We conclude that clinicians must interpret ABPs in the context of ASD, recognizing that they may differ substantially from those observed in typically developing peers.

## Data Availability

All data in the study is available from the NIMH Data Archive using a limited access data use certificate.

## Supplementary Material

### Differential Spearman Correlations

We let italicized letters like *A* denote a random variable and bold italicized letters like ***A*** denote sets of random variables. The *p* severity scores of the CASI-5 ***X*** and the *q* aberrant behaviors of the ABC ***Y*** have ordinal and incomparable scales. We thus assume that all random variables have been rank transformed and then normalized to mean zero standard deviation one. We then consider the linear correlation between each severity score *X*_*i*_ ∈ ***X*** and ***Y*** conditional on a diagnosis of ASD:

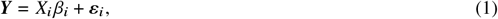

where *β*_*ij*_ corresponds to the standardized regression coefficient between *X*_*i*_ and *Y*_*j*_ ∈ ***Y***, equivalent to the Spearman correlation coefficient due to the rank standardization. The error terms *ε*_*i*_ in the above model satisfy 𝔼(*ε*_*i*_ | *X*_*i*_) = 0 and do not necessarily follow a Gaussian distribution. We focus on correlation rather than multivariate regression because we want to detect all predictive severity scores – not just a minimal set of predictive scores.

We likewise consider the Spearman correlations 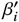 conditional on a diagnosis of TD via the following model:

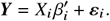

The *differential correlation* between ASD and TD then corresponds to *β* − *β*^′^.

### Orthonormal Components

We now wish to decompose ***Y*** into a set of orthogonal components, or approximately independent ABPs, for human interpretability. We focus on the following linear model obtained from an eigendecomposition of the correlation matrix of (unconditional) ***Y*** yielding *q* eigenvectors *V* and a diagonal matrix of *q* strictly positive eigenvalues Λ:

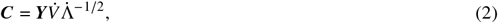

where 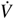 refers to *m* ≤ *q* of the eigenvectors with associated *m* eigenvalues 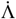. The variables ***C*** thus correspond to *m* orthonormal components of ***Y***.

Combining Equations (1) and (2) yields the following model from each *X*_*i*_ ∈ ***X*** to ***C*** conditional on a diagnosis of ASD:

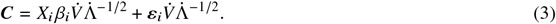

Now let 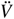 and 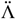 refer to the remaining *q* − *m* eigenvectors and their eigenvalues, respectively. We can then write:

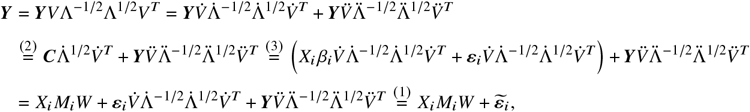

where 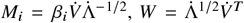 and 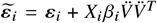. Numbers above the equality signs refer to equation numbers. The matrix *M*_*i*_ denotes the correlation from *X*_*i*_ to ***C***, and the matrix *W* denotes the effect sizes from ***C*** to ***Y***.

We likewise obtain the following model conditional on a diagnosis of TD:

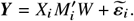

The differential correlation from *X*_*i*_ to ***C*** between ASD and TD then corresponds to 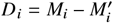. Let *D* now denote the differential correlation matrix, where *D*_*i*_ refers to the *i*^th^ row.

### Optimal Rotation

Note that we can rotate the orthonormal components ***C*** using the rotation matrix *R* and arrive at another set of orthonormal components ***C****R*. As a result, the transformation in Equation (2) is not unique. We thus now focus on specifying the best rotation *R*.

Let ℛ(*m*) denote the set of all *m* × *m* rotation matrices. We seek to identify severity scores that correlate with aberrant behaviors differently in ASD than in TD. As a result, we focus on the differential correlation matrix *D* and choose *R* ∈ ℛ(*m*) by solving the following optimization problem:

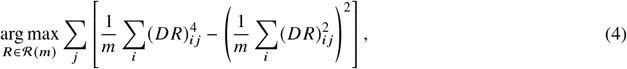

which is also known as the varimax rotation. This procedure ensures that only a few entries in each column of *DR* have large values, and the locations of the entries differ between the columns.^15, 26, 16^ Therefore, only a few maximally non-overlapping severity scores have a large differential correlation to each orthonormal combination of aberrant behaviors. If *R* denotes the solution to the above optimization problem, then the *optimal components* correspond to ***C***^∗^ = ***C****R*.

### Algorithm

We summarize Differentially Supervised Varimax (DSV) in Algorithm 1. In Line 1, DSV first ranks and standardizes both ***X*** and ***Y*** separately in ASD and in TD. DSV then computes the differential correlation matrix *D* in Line 2 and the matrix *W* from 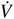 and 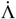 in Line 3. This allows the algorithm to perform the varimax rotation by solving Expression (4). The objective function only specifies the rotation up to sign and permutation indeterminancies.^27^ DSV therefore specifies the signs and permutations in Line 5 for use in downstream permutation testing.

#### Algorithm 1

Differentially Supervised Varimax (DSV)

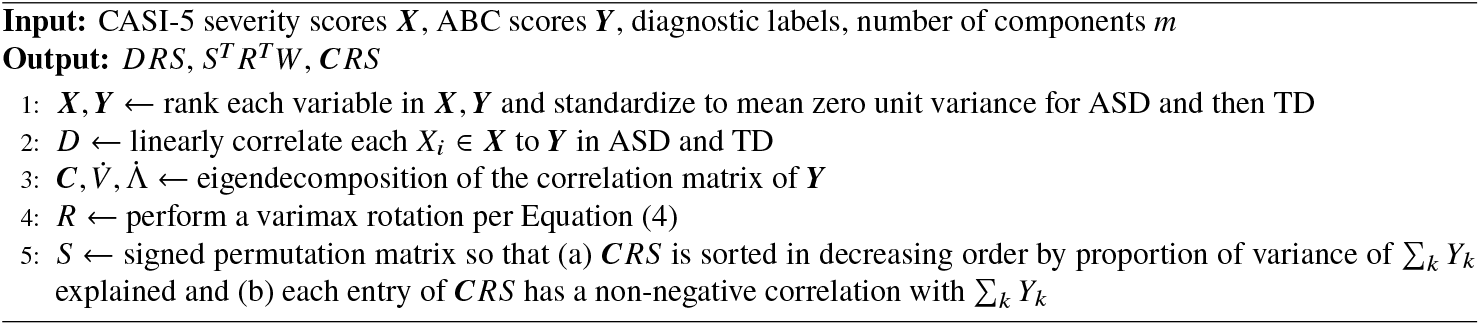

### Permutation Testing

We perform permutation testing by component and by severity-component as described below.

### By Component

We adopt the following null and alternative hypotheses:

- ℋ_0_ : (***X, Y***) **╨** *G*,
- ℋ_1_ : _*i*_ | (*DR*)_*i j*_ | > _*i*_ | (*DR*)_*i j*_ | _(***X***,***Y***) **╨***G*_.

where *G* corresponds to the diagnosis of either ASD or TD, and |(*DR*)_*ij*_| corresponds to the magnitude of the differential correlation from severity score *X*_*i*_ to optimal component 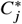. We permute the diagnosis, run the DSV algorithm and compute the *absolute sum* ∑_*i*_ |(*DR*)_*ij*_|. We repeat the process 100,000 times and then count the number of cases where the absolute sum in the permuted samples falls at or above the same absolute sum in the original samples. We repeat the process for each optimal component and thus compute *m p*-values in total. We estimated FDR-corrected *p*-values using the method described in [28]. We report the FDR-corrected *p*-values alongside the absolute sum statistic.

### By Severity-Component

We adopt the following hypotheses for each severity-component pair:

- ℋ_0_ : (***X, Y***) **╨** *G*,
- ℋ_1_ : |(*DR*)_*ij*_| > |(*DR*)_*ij*_| _(***X***,***Y***) **╨***G*_.

Permutation testing then proceeds similarly to the description above, where we count the number of cases where the magnitude |(*DR*)_*ij*_| in the permuted samples falls at or above the magnitude in the original samples. We repeated the test for each severity-component pair. This process results in a large number of *pm p*-values, so we correct the *p*-values by the FDR using the Storey method instead.^29^ We report the FDR-corrected *p*-values in Figure 3 and the differential correlation statistic (*DR*)_*i j*_ in each cell of Figure 4.

### Scree Plot

**Figure S1:**
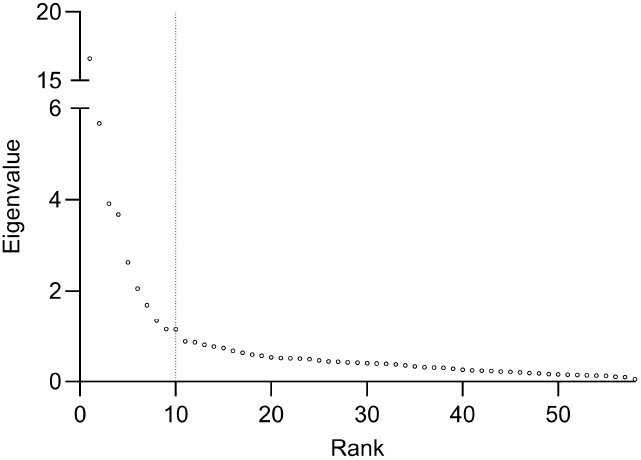
Scree plot displaying the ordered eigenvalues. We used *m* = 10 components due to the steep cliff observed in the scree plot after the tenth eigenvalue.

